# #ChronicPain: Automatic establishment of a chronic pain cohort from social media using machine learning for studying opioid-alternative therapies

**DOI:** 10.1101/2022.08.20.22279021

**Authors:** Sahithi Lakamana, Yuting Guo, Yao Ge, Abimbola Leslie, Omolola Okunromade, Yuan-Chi Yang, Elena Gonzalez-Polledo, Jeanmarie Perrone, Anne McKenzie-Brown, Abeed Sarker

## Abstract

Due to the high economic and public health burden of chronic pain, and the risk of public health consequences of opioid-based treatments, there is a need to identify effective alternative therapies. The evidence basis for many alternative therapies is weak or nonexistent. Social media presents a unique opportunity to gather large-scale knowledge about such therapies self-reported by sufferers themselves. We attempted to (i) verify the presence of largescale chronic pain-related chatter on Twitter, (ii) develop natural language processing (NLP) and machine learning for automatically detecting chronic pain sufferers, and (iii) identify the types of chronic pain-related information reported by them. We collected data from Twitter using chronic pain-related hashtags and keywords. We manually performed binary annotation of a sample of 4998 posts to indicate if they were self-reports of chronic pain experiences or not, and obtained inter-annotator agreement of 0.82 (Cohen’s kappa). We trained and evaluated several state-of-the-art transformer-based text classification models using the annotated data. The RoBERTa model outperformed all others (F1 score = 0.84; 95% CI: 0.80-0.89), and we used this model to classify a large number of unlabeled posts. We identified 22,795 self-reported chronic pain sufferers and collected their past posted data. Via manual and NLP-driven analyses, we found information about but not limited to alternative treatments, sufferers’ sentiments about treatments, side effects, and self-management strategies. Our social media-based approach will result in an automatically growing massive cohort over time, and the data can be leveraged to identify self-reported effective alternative therapies for diverse chronic pain types.

## INTRODUCTION

Between 5.5% to 33% of the world’s and over 30% of the United States (U.S.) adult population are estimated to suffer from chronic pain.^1–3^ The total financial burden of chronic pain in the U.S. is estimated to be between $560 and $635 billion per year.^4^ Opioid pain relievers are no longer considered first-line therapies because they are addictive and have been implicated in the epidemic of drug overdose-related deaths in the U.S.^3^ Due to the enormous financial and non-financial cost of chronic pain and its treatment with opioids, substantial recent research efforts have focused on identifying alternative treatments, including pharmacological, natural, and behavioral treatments.^5^ Currently, evidence is incomplete about the efficacies of many alternative therapies, including emerging therapies,^5–9^ the associations between types of pain and effective alternative management strategies, and the risks involved with the different strategies (*e*.*g*., long- and short-term side effects of opioid alternatives). It is difficult to establish the evidence and risk profiles of alternative therapies as they require the execution of studies in clinical settings, particularly trials. However, many alternative therapies have widespread use, and curating the knowledge from peoples’ experiences in a systematic manner may lead to the discovery of useful alternative therapies for targeted chronic pain types.

Many recent studies have leveraged social media big data for obtaining insights directly from patients about targeted health-related topics including, but not limited to, substance use,^10–13^ adverse drug reactions,^14–16^ and mental health.^17–19^ Over 220 million Americans (∼70%) use social media, and there is an abundance of health-related information on such platforms.^20,21^Recent advances in data science and machine learning have enabled researchers to extract complex information expressed in natural language, and have even made it possible to curate specialized cohorts from social media for conducting longitudinal studies based on data posted by the cohort members, such as breast cancer patients,^22^ pregnant people,^23^ and people who use substances.^24^ Social media thus present an attractive and underexplored opportunity for studying people suffering from chronic pain at low cost, unobtrusively, and at scale, provided that a chronic pain cohort can be built automatically from such sources, and longitudinal data from the cohort members can be detected.

In this paper, we take the first steps toward building a system that automatically detects self-reports of chronic pain from social media, specifically Twitter. Our study (i) verifies that reports of chronic pain are frequently posted on Twitter by sufferers themselves, (ii) such reports can be automatically detected via natural language processing (NLP) and supervised machine learning, and (iii) longitudinal data posted by self-reported sufferers of chronic pain contain a trove of knowledge about chronic pain therapies, including non-traditional therapies and self-management strategies, their efficacies, and risks. We focus this paper on the chronic pain cohort development process, leaving the tasks of mining longitudinal data and deriving potential causal associations as future work. The automatically-detected cohort will continue to grow over time and will enable us to study a massive cohort of people over many years.

## METHODS

This study was reviewed by the Institutional Review Board of Emory University and considered to be exempt (category 4; publicly available data). Figure 1 presents our data processing pipeline for this study. The overall study can be divided into 4 steps: (1) data collection, (2) manual annotation, (3) supervised classification, and (4) post-classification analyses. We now describe each of these steps.

**Figure 1:**
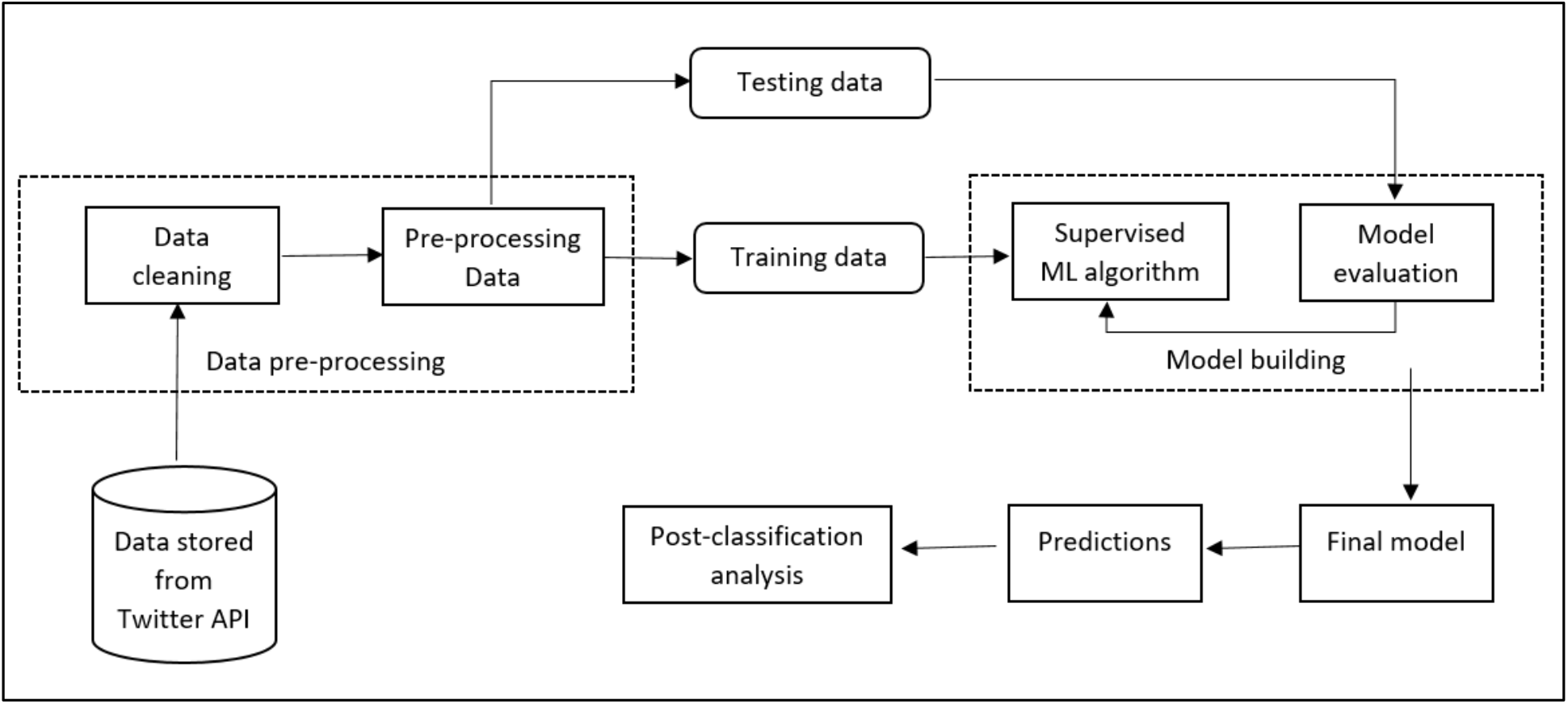
End-to-end data processing framework involving data collection/storage via the Twitter API, data preparation and preprocessing, training, evaluation, classification using the final model, and post-classification analyses.

### Data collection

We collected data through the Twitter academic streaming application programming interface (API).^25^ The API enables the collection of a sample of data in real-time based on keywords. We used the hashtag *‘#chronicpain*’ and the phrase ‘*chronic pain’*. For the phrases, we first collected tweets using the terms individually, and then applied a regular expression pattern to detect the exact phrase. We excluded retweets and tweets that did not meet the pattern.

### Annotation

We randomly selected 5000 posts from our entire collected set for manual annotation. The objective of the annotation was to determine if a post represented a personal experience or not.

Thus, the annotation was a binary labeling task indicating if a tweet represented a self-report (label: Y) or something other than a self-report (label: N). Label N represented tweets that were general information about chronic pain or those that could be about raising awareness or promotion. We conducted the annotation in three batches. In the first batch, we annotated a small sample of tweets (n=200) without an explicit guideline and we discussed the disagreements from this round of annotation to create a simple annotation guideline. We annotated a larger sample (n=1300; batch 2) following this guideline, and we further discussed disagreements and resolved them. We made minor updates to the annotation guideline based on these discussions. Since the disagreements were relatively low after the second round of annotations, we annotated the remaining tweets (batch 3).

Randomly selected subsets of the tweets were annotated by all three annotators. We used these overlapping annotations to compute inter-annotator agreements (IAA). Since the interpretation of the tweets can be subjective, it was important to capture the extent of human agreement as it represents a potential ceiling for any machine learning algorithm used to automate the process. We used the Cohen’s kappa measure to measure IAA. All disagreements were resolved by the last/senior author of the article (AS). The annotation guideline is available as supplementary material. Table 1 presents examples of tweets and their labels.

**Table 1.** Sample posts representing self-reported chronic pain (Y) and generic chronic pain-mentioning posts (N).^1^

| Post | Label |
| --- | --- |
| "A2. Being Black affects every aspect of my #ChronicPain experience. People don't believe my chronic pain experience. They perceive my pain as an over exaggeration. From doctors minimizing my chronic pain to assuming I can just ""endure"" it. | Y |
| Chronic pain will have you thinking the craziest thing. My hips are killing me so I thought it might be nice to not have legs... But then I realise my back and arms also hurts so then I thought what about being a floating head.. then I remembered migraines. #chronicpain | Y |
| 26 years old, 15 years of chronic pain and fatigue... finally diagnosed with Endometriosis #awareness #endometriosis #ChronicPain | Y |
| Good luck with that!! I have 3 herniated discs, sitting on nerves, spinal hypertrophy, chronic pain...diagnosed 7 months ago and my appt to see an NHS specialist THIS Oct has been cancelled Now on the waiting list No end in sight!!! Hope u have better luck #ChronicPain | Y |
| My chronic pain is gotten worse since my covid infection. I wake up with pain in my neck muscles every day now. | Y |
| "Here is a good paper from The Lancet looking at chronic pain: an update on burden, best practices, and new advances. Doesn't look like it's open access but you might be able to access it from your institution. [link] | N |
| 20 Tips: Making the Best of 20 Years of Chronic Pain and Illness [link] #chronicpain | N |
| "Plant-derived compound may help treat chronic pain. #chronicpain #CRP [LINK]" | N |
| "Chronic pain and low oxalate diet? Some people find a huge amount of relief from trying a low oxalate diet when they have chronic pain. Have you tried it? Let me know #chronicpain #spoonie #chronicillness #cfs #fibro" | N |

### Supervised classification

We divided the annotated data into 3 sets—60% for training, 20% for validation, and 20% for testing/evaluation. Transformer-based approaches that use large pre-trained language models such as BERT are currently state-of-the-art for text classification, both in and outside of the medical domain. We, therefore, fine-tuned and evaluated several transformer-based classifiers. The following is an outline of the classifiers we used:

1. RoBERTa: Transformer-based model popular for its training on big batches and long sequences.^26^
2. SciBERT: Model is trained on a large corpus of scientific data and with the same model structure as BERT.^27^
3. BioClinicalBERT: This was modelled from BioBERT (BioBERT-Base v1.0 + PubMed 200K + PMC 270K) and was trained on MIMIC III notes.^28^
4. BERTweet: Large scale pre-trained model for English tweets.^29^
5. BioBERT: This is specifically trained for bio-medical text and is widely used for biomedical text mining.^30^

We compared the performances of the classifiers based on the F_1_ score for the Y class. The F_1_ score is the harmonic mean of precision and recall. We focused our evaluation on the Y class since that is our class of interest. We also report overall classifier accuracy, but this metric is primarily driven by the majority (N) class. We computed the 95% confidence intervals for the Y class F_1_ scores using the bootstrap resampling technique.^31^ In this method, the F_1_ score is calculated 1000 times for randomly selected (with replacement) training sets. Out of these, 25^th^ and 975^th^ values are considered as lower bound and upper bound values at 95% confidence interval.

### Post-classification analyses

We used the best-performing classifier from the previous subsection to classify unlabeled posts collected from January 2021 to March 2022. Then, for a sample of subscribers whose posts were classified to be self-reports of chronic pain (Y), we collected all their past posts available via the API. The Twitter API allows the collection of approximately 3200 past public posts by each subscriber. Thus, for many subscribers—members of our chronic pain cohort—this enabled us to obtain multiple years of posts. We were only able to collect this for a subset of the subscribers due to the API limitation of 10 million post collection per month. Finally, we semi-automatically analyzed samples of the cohort posts to assess the presence and type of chronic pain-related information. The semi-automatic analysis involved using a lexicon-based fuzzy matching approach^32^ to detect potential treatments mentioned, including pharmaceuticals such as opioid pain relievers (*e*.*g*., oxycodone, hydrocodone) and non-standard ones such as behavioral therapies. We performed a number of analyses involving the posts that mentioned specific therapies and also the past posts collected from the subscribers. We outline these below.

#### Therapy-related posts analysis

We extracted posts that were detected to mention at least one therapy and compared their distributions. We also drew a small sample of posts mentioning therapies and conducted a sentiment analysis. Sentiment analysis was performed to obtain an estimate of how subscriber sentiments were associated with each therapy, if at all, and if the data held potentially differentiable therapy-specific sentiments. Two co-authors (LA, OL) manually reviewed each post and determined whether the sentiment expressed in relation to the therapy was (i) positive, (ii) neutral, or (iii) negative. We compared the overall distributions of the mentioned therapies in our cohort data and also the distributions of sentiment associated with each therapy.

#### Cohort post content analysis

For Twitter subscribers who mentioned more than 3 chronic pain- or treatment-related posts, we also reviewed samples of their unlabeled tweets to identify other information relevant to chronic pain research and the topics of discussion. In-depth analyses of the types of information posted by the cohort and their quantification were considered to be outside the scope of this study. Instead, we simply focused on characterizing the types of relevant information present and verifying their contents for future analyses.

## RESULTS

### Data and annotation

Of 5000 posts selected for annotation, two were excluded due to issues with text encoding, leading to the annotation of 4998 posts in total. 550 posts were at least double annotated. Pair-wise inter-annotator agreement (IAA) among 3 annotators was *k*=0.82 (Cohen’s kappa^33^), which can be considered to be *almost-perfect agreement*.^34^ 719 (14.4%) were self-reports of chronic pain while the rest (85.6%) were not. The class distribution was thus slightly imbalanced. This was unsurprising since our past studies have found similar imbalances.^35,36^

### Classification performance

Table 2 presents the results of our automatic classification experiments. The table shows the overall accuracy of the classifiers, the F1 scores for the positive class and the 95% confidence intervals. The RoBERTa classifier achieved the highest F1 score among all the classifiers. For the best-performing classifier, we conducted ablation experiments using 10% subsets of the original training data. Figure 2 shows, and as is typically seen from such experiments, that the performance of the classifier increases approximately logarithmically as more training data is added. The best-fit logarithmic trendline suggests that F1 scores of 0.87 would require us to increase the annotated dataset by three times. Accuracy scores are relatively unchanged with training data size since this value is primarily driven by the majority negative class.

**Table 2.** Classifiers, their overall accuracies, F1 scores, and 95% Confidence Intervals (CIs) for the F1 scores.

| Classifier | Accuracy | $F_1$ score (Y class) | 95% CI of $F_1$ score |
| --- | --- | --- | --- |
| RoBERTa | 0.95 | 0.84 | 0.80-0.89 |
| SciBERT | 0.93 | 0.79 | 0.74-0.84 |
| BioClinicalBERT | 0.95 | 0.82 | 0.76-0.86 |
| BERTweet | 0.95 | 0.83 | 0.78-0.87 |
| BioBERT | 0.95 | 0.82 | 0.76-0.86 |
| Clinical_KB_BERT | 0.94 | 0.80 | 0.74-0.84 |

**Figure 2:**
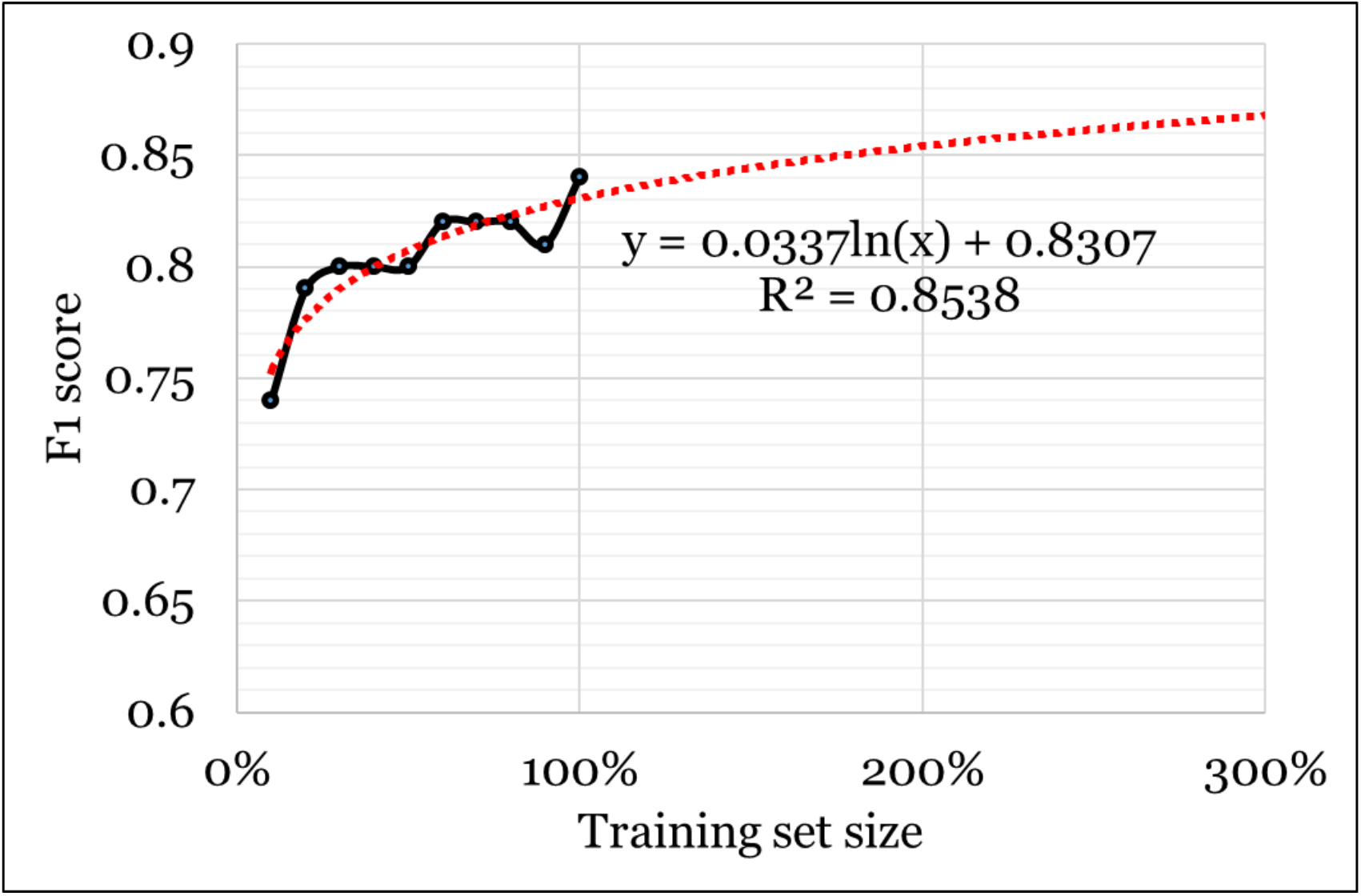
Classifier F_1_ scores or the positive (Y) class at different training set sizes and projected scores based on a logarithmic trendline for the RoBERTa model.

### Post classification analyses

The application of our classifier on the unlabeled posts resulted in the identification of 41,262 self-reports of chronic pain from 22,795 subscribers. Collecting their past data resulted in 3,461,619 posts. We performed the post classification analyses on these posts, and we described the findings in the following subsections.

#### Therapy-related post analysis

Table 3 presents the therapies we discovered from the cohort-posted data and their distributions. The therapies we discovered include prescription medications such as opioids, non-prescription pharmacological substances such as cannabidiol (CBD) and cannabis, physical therapies such as massage and chiropractic, and behavioral/physical therapies such as meditation and yoga. We even discovered many examples of relatively unexplored therapies such as music therapy, aromatherapy, and guided imagery. Cannabis was the most commonly mentioned substance in the cohort timelines, although our manual review suggested that many of the posts were about recreational use rather than their use for treating chronic pain. A substantial number of tweets, however, did describe the use of cannabis-related products for treating chronic pain. Chiropractic was the most commonly mentioned physical therapy. Table 4 presents examples of tweets mentioning therapies, the therapies mentioned, and their categories. As illustrated in the table, posts often describe self-management strategies for chronic pain, pharmacological substances, and their efficacies, comparisons between different pain management strategies, and adverse effects of therapies (*e*.*g*., opioid pain relievers) including long-term impacts (*e*.*g*., addiction).

**Table 3.** Therapies mentioned by our chronic pain cohort and their distributions in the longitudinal cohort data.

| **Treatment** | **Number of posts** |
| --- | --- |
| delta-8/weed/marijuana/cannabis/THC | 21,829 |
| CBD | 6,312 |
| Chiropractic | 4,765 |
| Yoga | 3,162 |
| Massage | 3,148 |
| Meditation | 2,244 |
| Acupuncture | 1,919 |
| Dietary | 800 |
| Relaxation | 756 |
| Hypnosis | 483 |
| Morphine | 338 |
| Buprenorphine | 332 |
| Methadone | 214 |
| Tramadol | 178 |
| Aquatic | 166 |
| Tai Chi | 146 |
| Aromatherapy | 128 |
| Music Therapy | 120 |
| Oxycontin | 108 |
| Biofeedback | 71 |
| Hydrocodone | 56 |
| Guided Imagery | 34 |

**Table 4.** Sample posts extracted from the cohort timelines that mentioned at least one therapy for chronic pain.

| **Post** | **Therap[y\|ies]** | **Therapy Type(s)** |
| --- | --- | --- |
| stopped my yoga for a couple of weeks and now my back is killing me again | Yoga | Behavioral/physical |
| I have been reducing my opioid dosage after over a decade of taking them for chronic pain (jaw/face) and struggling with them greatly at times. TODAY I AM OFFICIALLY OFF MORPHINE! No more Fentanyl, OxyContin etc. either! Today I am proud of myself! | Opioid<br>Morphine<br>Fentanyl<br>Oxycontin | Pharmacological (prescription) |
| First time I was prescribed Oxycontin, my doctor had me watch a video about how if you truly had chronic pain, you could not become addicted to Oxycontin.<br>I became addicted to Oxycontin. | Oxycontin | Pharmacological (prescription) |
| Then I was told I didn't have *real* pain, because if I did, I couldn't have been addicted. |  |  |
| Exercise and diet helped my neck pain way more than those pain meds | Pain meds<br>Exercise<br>Diet | Pharmacological<br>and<br>behavioral/physical |
| If it weren't for cannabis, I would still be addicted to opioids and my pain would never get better | Cannabis<br>Opioids | Pharmacological |
| 'm a chronic pain patient. I used to take 3 big gun oxycontin a day and overuse my scripts. Today, I use cannabis which isn't chemically addictive and has impressive medical benefits for those genuinely sick and standing in need | Oxycontin<br>Cannabis | Pharmacological |
| Have you tried acupuncture? My chiro couldn't help so I tried it & it worked wonders!! J | Acupuncture<br>Chiropractic | Behavioral/physical |
| Now I am trying CBD... Will let ya know if it helps in any way | Cannabidiol | Pharmacological |

#### Sentiment Analysis

Sentiment Analysis is performed to estimate the polarity distribution of feelings associated with the chronic pain experienced by the cohort members who also mention specific treatment keywords. 600 posts were manually annotated in total with an IAA of 0.88 (Cohen’s kappa). Figure 3 presents the sentiment distributions for different types of therapies. The figure shows that most of the users who mentioned “*meditation*”, “*Guided imagery*”, and “*Tai Chi*” in their posts, also showed more positive emotions, while users who mentioned “*hydrocodone*” and “*tramadol*” show negative emotion compared to others.

**Figure 3:**
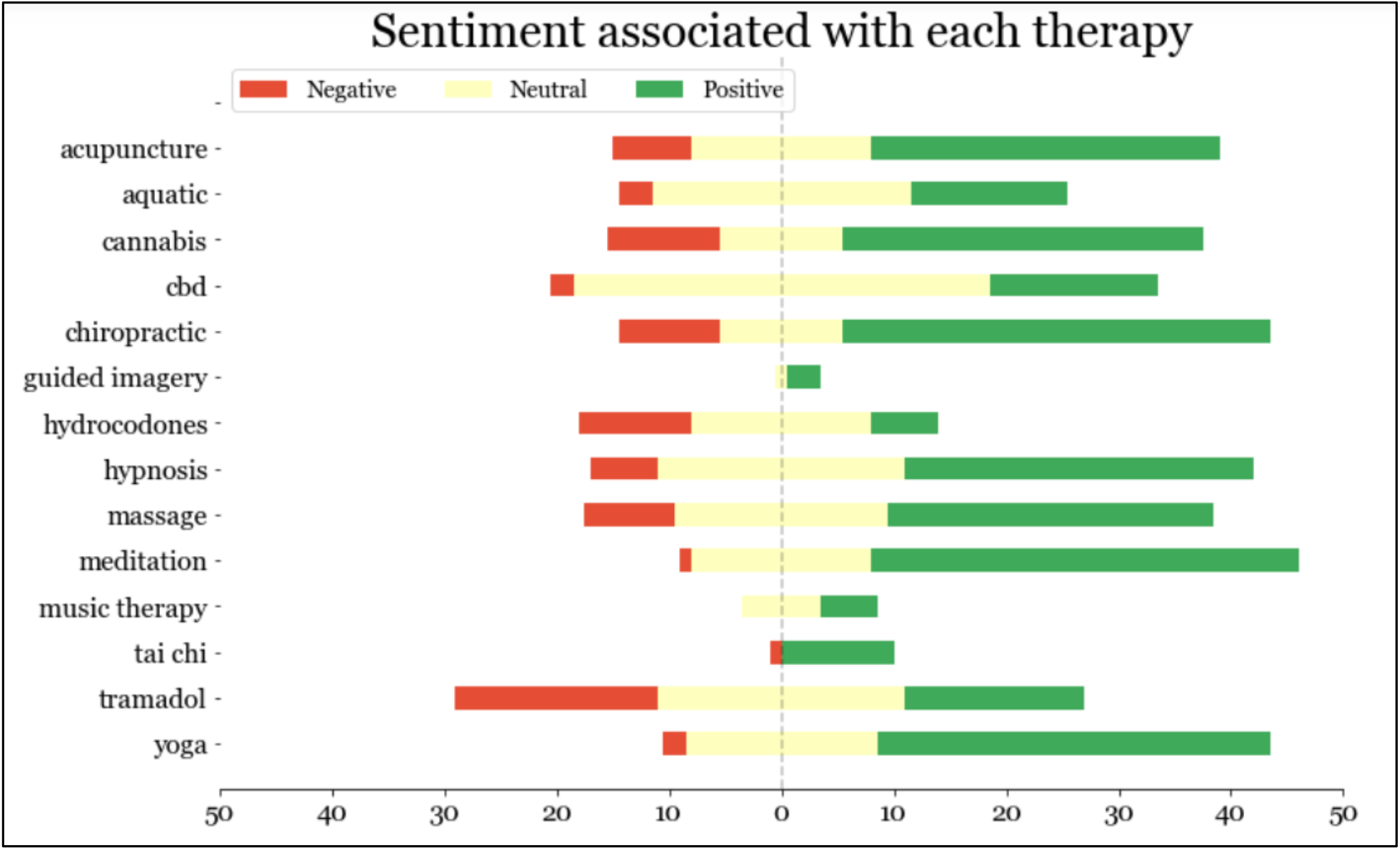
Distribution of manually annotated sentiments (positive, negative and neutral) for different chronic pain therapies, as discussed by the cohort members.

#### Cohort post content analysis

Table 5 presents examples of posts from the longitudinal cohort data that present chronic pain related information. The types of relevant information posted by the cohort include but are not limited to: descriptions of pain, therapies, impacts of therapies, self-management strategies, social support, and its impact, mental health, treatment access-related information, and questions about chronic pain. The presence of such wide-ranging information suggests that publicly available data from this cohort may be invaluable in conducting long-term chronic pain-related studies. We further discuss the utilities of this data source and our data curation strategy in the following section.

**Table 5.**
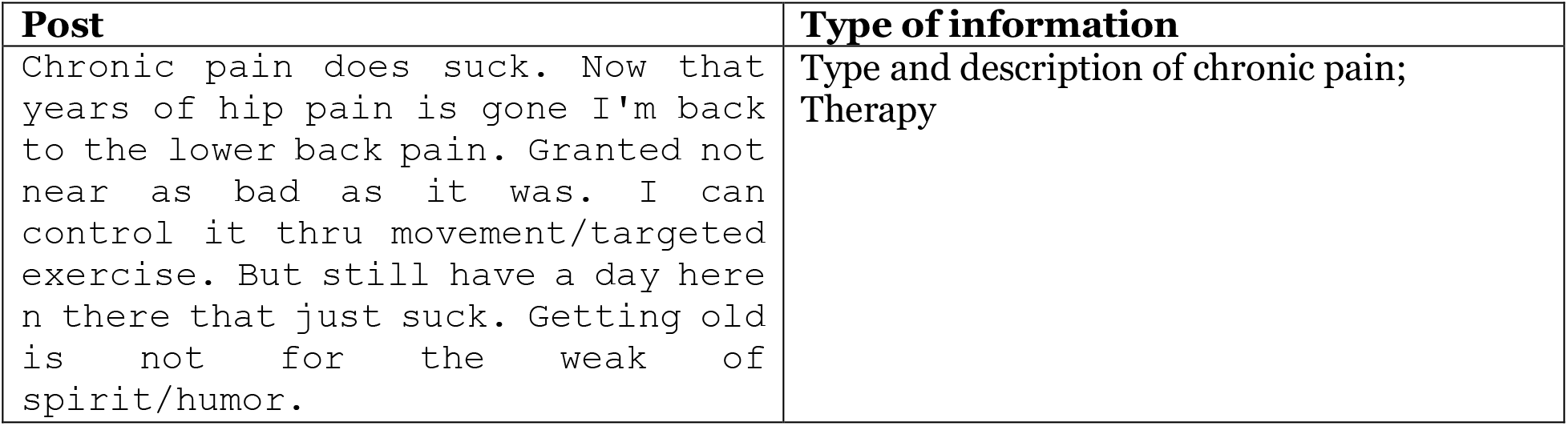
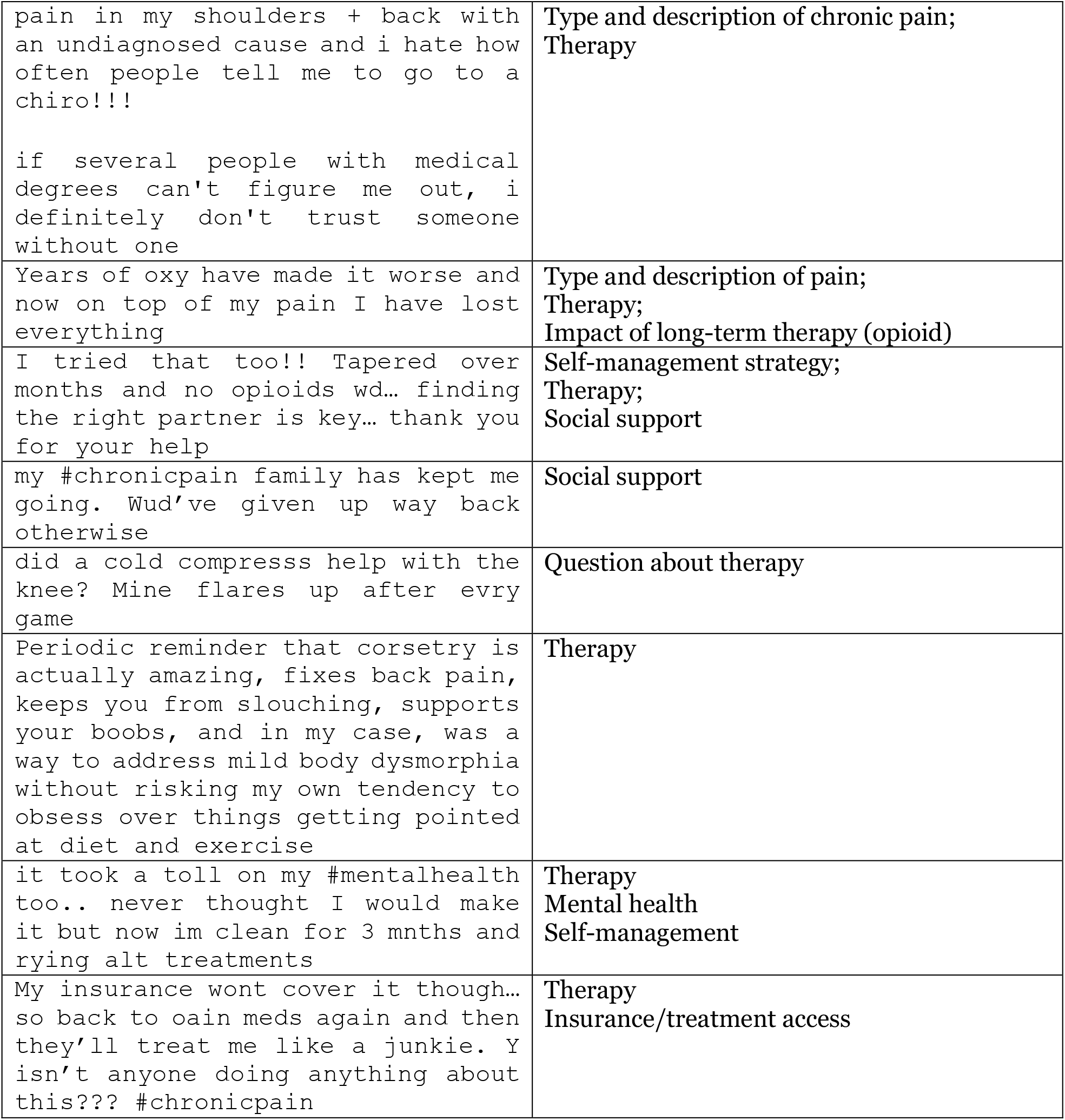
Sample posts relevant to chronic pain detected within the public timelines of our automatically-built cohort. The posts present a plethora of information including the types and descriptions of chronic pain, therapies, social support, questions about therapy and chronic pain, self-management strategies, mental health, and treatment access (including health insurance coverage).

| Post | Type of information |
| --- | --- |
| Chronic pain does suck. Now that years of hip pain is gone I'm back to the lower back pain. Granted not near as bad as it was. I can control it thru movement/targeted exercise. But still have a day here n there that just suck. Getting old is not for the weak of spirit/humor. | Type and description of chronic pain; Therapy |
| pain in my shoulders + back with an undiagnosed cause and i hate how often people tell me to go to a chiro!!!<br>if several people with medical degrees can't figure me out, i definitely don't trust someone without one | Type and description of chronic pain;<br>Therapy |
| Years of oxy have made it worse and now on top of my pain I have lost everything | Type and description of pain;<br>Therapy;<br>Impact of long-term therapy (opioid) |
| I tried that too!! Tapered over months and no opioids wd... finding the right partner is key... thank you for your help | Self-management strategy;<br>Therapy;<br>Social support |
| my #chronicpain family has kept me going. Wud've given up way back otherwise | Social support |
| did a cold compresss help with the knee? Mine flares up after evry game | Question about therapy |
| Periodic reminder that corsetry is actually amazing, fixes back pain, keeps you from slouching, supports your boobs, and in my case, was a way to address mild body dysmorphia without risking my own tendency to obsess over things getting pointed at diet and exercise | Therapy |
| it took a toll on my #mentalhealth too.. never thought I would make it but now im clean for 3 mnths and rying alt treatments | Therapy<br>Mental health<br>Self-management |
| My insurance wont cover it though... so back to oain meds again and then they'll treat me like a junkie. Y isn't anyone doing anything about this??? #chronicpain | Therapy<br>Insurance/treatment access |

## DICUSSION

Our goals in this study were to verify that social media, particularly Twitter, contains chatter about chronic pain posted directly by people suffering from it, manually annotate a sample of posts mentioning chronic pain to indicate if they represented self-reports or not, training and evaluating several supervised classification algorithms to automatically detect self-reports of chronic pain, building a cohort of chronic pain sufferers over Twitter for long-term, longitudinal analyses, and semi-automatically analyzing a sample of posts from the cohort to determine the presence of chronic pain relevant information that can be studied in detail in the future. Our study verifies that social networks such as Twitter contain valuable information about chronic pain, posted by sufferers themselves. However, while such valuable information does exist, separating it from the massive volume of data that is constantly generated on Twitter is a challenging task. It is impossible to continuously curate such data manually. Our proposed methods automated the process of (i) detecting chronic pain related posts automatically in real-time via the Twitter API, and (ii) identifying posts that represented self-reports (*i*.*e*., patients describing their own experiences). This strategy of automatic data collection and cohort curation will be able to establish a massive cohort over time, and will enable us to study multiple years of data, including longitudinal data from the entire cohort and its subsets. This innovative strategy has additional advantages— (i) it allows us to include people who may not be reachable via traditional settings, such as hospital-based settings (*e*.*g*., because of lack of health insurance); (ii) it is very cost effective and is unobtrusive, and (iii) it is able to continuously grow the cohort, thus enabling us to gather big data for long-term research.

In the rest of this section, we first outline some related work in order to put our contributions into context. We then verify the utility of automatic classification approaches for monitoring and studying chronic pain. Finally, we present a brief error analysis to identify essential future improvements and we discuss some of the limitations of this study.

### Related work

Several past studies have demonstrated the utility of social media data for chronic pain-related research. Many early studies focused on studying the effects of social media for chronic disease management, rather than the use of it to study chronic pain specifically.^37,38^ Through a global online survey conducted partially via social media, Merolli et al.^38^ concluded that areas of research that warranted attention included the ability to (i) filter information and guide people to pertinent information, (ii) connect sufferers of chronic pain (*i*.*e*., cohort members), and (iii) explore relationships between the therapeutic affordances of social media and health outcomes. In a separate, questionnaire-based study, Ressler et al., found that posting about chronic pain online may decrease the sense of isolation and increase a sense of purpose.^39^ Similar findings were reported by Tsai et al. in a more recent study.^40^ Gonzalez-Polledo et al. studied social media (Tumblr and Flickr) and chronic pain from the perspective of digital anthropology, characterized chronic pain narratives over these platforms, and presented a typology of chronic pain expressions.^40^

Works most closely related to ours are those by Sendra and Farre and Mullins et al. ^39,41^. In the former study, the authors analyzed a small sample of data (n=350) and concluded that social media is changing the way patients live with their chronic pain and care providers could benefit from paying attention to self-reported information by these individuals. Mullins et al. collected a small sample of data from Twitter and performed NLP-driven analysis of discussion topics, sentiments and advice provided. The study concluded that the pain-related discussions on the platform can enrich our understanding of the chronic pain experience. Our study builds on these and proposes a sustainable infrastructure, driven by NLP and machine learning, that can automatically curate knowledge from a chronic pain cohort over time. No prior work has proposed as thorough an approach. Crucially, our study goes beyond relying on post-level information by focusing on building a chronic pain cohort and collecting data solely from this cohort. This ensures that the texts included in the analyses are more likely to be related to personal experiences of chronic pain rather than general discussions about the topic. Our past efforts to build targeted cohorts have led to the creation of massive, multi-year datasets involving hundreds of thousands of cohort members (*e*.*g*., people who use substances^42^ and pregnant people^23^).

### Utility of social media

Our findings illustrate that social media is a rich source of information for studying chronic pain. Chronic pain-related discussions are common on Twitter, but this resource has thus far been underutilized in both epidemiological and interventional research. A prime reason for this is the difficulty of handling such massive data and processing complex language. Our proposed machine learning and NLP methods have the potential of overcoming the barriers leading to the underutilization of such an effective source of information. Successful deployment of this pipeline is likely to increase the utility of social media in chronic pain research. Our study also found that the Twitter chronic pain cohort discusses a variety of pain-related topics including but not limited to therapies, addiction, mental health, and support. Conducting more detailed analyses of texts associated with these topics may lead to important breakthroughs in chronic pain research. For example, analyses of the opioid alternatives discussed by subscribers can generate hypotheses about their efficacies, which can be tested in future research.

Studying a social media-based cohort may also reveal information about factors impacting the sufferers of chronic pain that may not be available from any other source. These may include, for example, their social support (e.g., number of followers, number of interactions with a given post) and its influence (if any) on the quality of life of chronic pain patients. There is also the potential of going beyond epidemiological studies over social media and conducting interventional studies.

### Error analysis

We conducted manual analysis on the misclassified tweets to identify patterns of errors. We focused on both false positives (classified erroneously as self-report) false negatives (classified as not self-report). Under the former category, we found that most posts were misclassified when users intended to share the experiences of the people around them, not their personal experiences. One such post is, *“today is mother’s day and it’s not an easy day. although my mom is still biologically alive, it feels like i’ve lost her to her chronic pain years ago. they say to treasure every moment but it’s not always easy”*. Under the latter category where model fails to classify as self-reports, we found that one common reason for misclassification was due to the expressions being implicit, making it difficult for the machine learning algorithms to capture their true meanings. One such example is, **“***music therapy for <hashtag> chronicpain. thinking of ways to make movement enjoyable might involve <hashtag> music <hashtag> dance. sometimes I’ll just chuck on some of my favorites and dance, mindfully. i always feel a million bucks after as well as more connected with my body*.*”* Another reason for misclassification in this category was determined to be incomplete information/partial information. For instance, “*<hashtag> finally starting to ease, been trying to focus my mind on other things to get my brain to turn down the chronic <hashtag> pain it’s at <number> out of <number>”*. While for a human reviewer the post has enough context, it could not be picked up by the machine learning classifier.

## Limitations

The most important limitation of the current method is perhaps the performance of the machine learning classifier. The F1 score for the positive class is relatively low at 0.84. Since most of the posts are not self-reports of chronic pain, the dataset is highly *imbalanced*—the negative class comprises most of the dataset. Improving performance over the minority class is a commonly addressed problem in text classification. The most common and effective strategy is to annotate more data manually so that the system can better generalize the features of the minority class instances. Our post-classification analyses show that the performance of the best classifier does not fully plateau with the data that is currently available for training, meaning that further annotation of data is likely to improve performance albeit only slightly.

Another major limitation is associated with the platform—social media subscribers tend to be younger, and more tech savvy compared to the general population. Thus, Twitter is not fully reflective of the U.S. or global population. Furthermore, only information publicly shared by our cohort is available for analyses. Information never shared by a subscriber will thus be missing. The former limitation is not unique to Twitter or social media—no data source is bias-free. Social media, however, is perhaps the platform with the best reach of any. As for the latter limitation, it is possible that certain population groups will share more information over social media compared to others (*e*.*g*., women have been found to share more chronic pain related information compared to men), and we did not adjust for that.

## Conclusion and future directions

Our study verified that (i) social media, specifically Twitter, contains a trove of information about chronic pain, directly posted by sufferers themselves, and (ii) this information can be automatically mined via NLP and machine learning methods to study targeted topics. This is the first such elaborate pipeline that attempts to curate knowledge about chronic pain from patient-generated social media data. The performances of our automatic methods are promising, despite the relatively low volume of manually annotated data. There is room for improvement in the machine learning model, and we will target such improvements in future work. Specifically, we will annotate more data and experiment with more sophisticated machine learning strategies (*e*.*g*., ensembling or fusion-based classification). Further improving performance will increase the quality of our chronic pain cohort. Importantly, the collection of cohort members via classification and their posts over time (*i*.*e*., the pipeline depicted in Figure 1) is fully automated, and so the cohort and the data will continue to grow over time. This will lead to the creation of an unprecedented resource for conducting long-term studies on the topic.

## Data Availability

All data produced in the present study are available upon reasonable request to the authors

## ACKNOWLEDGMENTS

Research reported in this publication was supported in part by the National Institute on Drug Abuse (NIDA) of the National Institutes of Health (NIH) under award number R01DA046619. The content is solely the responsibility of the authors and does not necessarily represent the official views of the NIH/NIDA.

The authors have no conflict of interest to declare.

## Footnotes

1 Posts were modified to preserve anonymity of the posters.

